# Extensive SARS-CoV-2 testing reveals BA.1/BA.2 asymptomatic rates and underreporting in school children

**DOI:** 10.1101/2023.01.15.23284579

**Authors:** Maria M. Martignoni, Zahra Mohammadi, JC Loredo-Osti, Amy Hurford

**Author notes:** **Ethics approval:** The study was approved by the Health Research Ethics Board (HREB) at the meeting held on January 27, 2022 (Reference number: 2022.013). **Consent to participate:** Participation was voluntary, and a consent was required before the survey questions were released. The consent form was approved by the Health Research Ethics Board (HREB). **Consent for publication:** The consent form and the ethic approval application specifies that results will be made public, and therefore consent for publication was provided by approval of the consent form. Consent form and signed ethic approval are attached. Availability of data and material: Data is publicly available at https://github.com/nanomaria/RAT_january. **Code availability:** The code used for the analysis is publicaly available at https://github.com/nanomaria/RAT_january. **Author contributions:** MM wrote the manuscript and analyzed the data. AH and J-LO provided guidance on the analysis. ZM provided constructive feedback on the preliminary analysis and results. AH designed and publicized the study, and obtained all necessary approvals. **Funding:** This study was funded by subgrants from the Emerging Infectious Disease Modelling Initiative groups of the Canadian Network for Infectious Disease Modelling, Mathematics for Public Health, and the One Health Modelling Network for Emerging Infectious Diseases to AH.

## Abstract

Case underreporting during the COVID-19 pandemic has been a major challenge to the planning and evaluation of public health responses. Inconsistent underreporting can undermine effective risk assessment due to high uncertainty in predicted future scenarios. Underreporting rates have been particularly high among children and youth, given that asymptomatic school children were often considered a less vulnerable population. In January 2022, the Canadian province of Newfoundland and Labrador (NL) was experiencing an Omicron variant outbreak (BA.1/BA.2 subvariants) and public health officials recommended that all students returning to elementary, junior high, and high schools (∼59,452 students) complete two rapid antigen tests (RATs) to be performed three days apart. To estimate the prevalence of SARS-CoV-2, we asked parents and guardians to report the results of the RATs completed by K-12 students using an online survey, and to specify the students’ school level and if students with positive RAT results had symptoms. When comparing the survey responses with the number of cases and tests reported by the NL testing system, we found that 1 out of every 4.3 (3.1-5.3) positive households were captured by provincial case count, with 5.1% positivity estimated from the RAT results, and 1.2% positivity reported by the provincial testing system. The survey data indicate that a higher percentage of SARS-CoV-2 cases were found in elementary schools, with 62.9% of positive cases (95% CI: 44.3%, 83.0%) reported from elementary school students, and the remaining 37.1% (95% CI: 22.7%, 52.9%) reported from junior high and high school students. Asymptomatic infections were 59.8% of the positive cases, with no significant difference between asymptomatic rates in elementary schools (60.8%) or in junior high and high schools (58.1%). Given the low survey participation rate (3.5%), our results may suffer from sample selection biases, and should be interpreted with caution. Nonetheless, our estimate of the underreporting ratio is consistent with ratios calculated from serology data, and our study provides insights into infection prevalence and asymptomatic infections in school children, a currently understudied population.

**We declare that:**

- This manuscript is original and is not a violation or infringement of any existing copyright or license
- The manuscript is not under consideration elsewhere
- All authors meet the definition of authorship as set out by the International Committee of Medical Journal Editors (ICMJE)
- Permission has been obtained from the copyright holder(s) if indicated, for the use of any third-party textual, graphic, artistic or other material

## 1 Introduction

During a pandemic, surveillance is essential for forecasting health care demand and to inform public health decisions. Infection underreporting and inadequate surveillance can lead to unreliable predictions, undermining effective risk assessment [61]. Underreporting of Severe Acute Respiratory Syndrome Coronavirus 2 (SARS-CoV-2), causing COVID-19, has been a major challenge to the analysis of epidemiological data and the implementation of preventive and control measures [37]. During the pandemic, COVID-19 prevalence has been inconsistently underreported for different reasons, such as challenges in maintaining a high testing capacity [27], discouraged testing of non-symptomatic individuals [11], and many mild or asymptomatic infections, particularly in children and youth [33]. Challenges to providing accurate COVID-19 case counts have increased with the establishment of more transmissible variants, leading to faster, and at times, uncontrolled infection spread [58], with the promotion of self-testing alongside no requirement that these results be reported [30, 64], and with increased vaccination coverage, decreasing the likelihood of severe outcomes and the need to seek health care [50]. All these factors have led to inconsistent variation of case reporting over time, challenging epidemic forecasting.

The Omicron variant of SARS-CoV-2 (formerly BA.1 or B.1.1.529.1, with sister lineage BA.2 or B.1.1.529.2), was first detected in South Africa on November 8, 2021, and declared a variant of concern by the World Health Organization on November 28, 2021 [15]. The Omicron variant spread extremely rapidly around the world. In Canada, the first Omicron variant case was reported in Ontario on November 28, 2021 [16], and in Newfoundland and Labrador (NL), the first Omicron variant case was reported on December 15, 2022 [18]. Before the spread of the Omicron variant, there was limited spread of SARS-CoV-2 in the NL community [36]. Until this time, NL had implemented a containment strategy, consistent with an elimination (or zero-COVID) strategy [26, 35]. This containment strategy limited SARS-CoV-2 spread through strict border control, contact tracing and self-isolation requirements, and non-pharmaceutical interventions aimed to end community transmission whenever outbreaks occurred [7].

When Omicron variant infections began spreading in the community, NL reported its highest COVID-19 case counts since the beginning of the pandemic. On January 17, 239 new cases were reported [11], which is 0.45% of the provincial population. After January 17, the province no longer publicly reported cases by age group. However, until then 19.7% of the reported cases were in the *<*20 age group, and a more detailed overview over the epidemiological situation in NL is provided in Appendix D and in [8].

With the Omicron variant’s higher transmissibility, its potential to escape the human immune response (meaning that vaccinated individuals and individuals that have already had COVID-19 may be susceptible for reinfection [40]), and, at the time, unknown health risks, these high case counts raised concerns of health care capacity overload. NL elementary (grades K-6), junior high (grades 6-7), and high schools (grades 8-12) closed early for Winter break on December 20, 2021 [20]. To reduce infection spread and protect the health care system, the return to in-person teaching for these students was postponed to January 25, 2022 [19].

In addition to the delayed return to school, public health officials strongly recommended that K-12 students (∼59,452 individuals) complete Rapid Antigen Tests (RATs; [11, 14]). The Department of Health and Community Services distributed BTNX Rapid Response COVID-19 antigen test kits to schools, and the schools distributed the kits to students. A first RAT was to be completed on January 22, three days before the return to in-person school. Students testing negative were asked to complete another test the morning of January 25 just before returning to school. Students that recorded positive test results were to self-isolate for 7 to 10 days, depending on their vaccination status [3]. At the time, 89.1% of the NL population aged 5 years and older, and 85.7% of the total population, were fully-vaccinated (defined as two doses [21]). The reason that school students were to complete these RATs was to ‘reduce the risk of someone attending school while infected’ [7]. There was no requirement to report these RAT results, but positive results could be submitted using the provincial COVID Assessment and Reporting Tool.

The wide distribution of RATs throughout the province, and the recommendation from public health officials for school students to complete these RATs on specific dates, allowed us to study the underreporting of the Omicron variant (BA.1/BA.2 subvariant), and infection prevalence in NL school students. Between February 3 and February 19, 2022, we deployed an internet survey that enabled parents and guardians to voluntarily report the number of positive and negative results for RATs completed by school students (grades K-6 or 7-12) on January 22 and 25. Our survey was unrelated to the provincial COVID Assessment and Reporting tool. Parents were asked to specify whether positive cases were symptomatic or asymptomatic, the Forward Sortation Area (FSA) and Regional Health Authority (RHA) where the tests were completed, and results for students in one household were to be reported together (see Appendix A).

The recommendation for school children to complete these RATs first occurred on January 13, and February 3 was the earliest we could begin the internet survey due to the time it took to obtain the necessary approvals. To ensure informed consent, as many students were under 19 (the age of majority in NL [22]), parents and guardians were asked to report the RAT results, but the reported test results were only for K-12 students. FSA is the first 3 digits of a postal code, and we asked participants to report their FSA so we could determine if spatially adjacent infection spread was occurring, and if there was substantial variation in infection prevalence within and between RHAs. We requested that results be reported together for one household because the Omicron variant is highly transmissible within a household [25], and therefore household positivity (rather than individual positivity) is a more reliable measure of prevalence, given that test results from individual students living in the same household are not independent. Furthermore, to estimate underreporting, we compared the results of the RATs with COVID-19 cases reported by the provincial testing system. This comparison was made at the household level because beginning January 24, 2022, it was stated that household members of COVID-19 cases in NL should not undergo testing at the provincial testing sites (PA2 11; see also, Appendix E).

Until 2021, COVID-19 testing in Canada occurred mostly for symptomatic individuals, and testing of asymptomatic individuals occurred in vulnerable populations, which included the elderly, residents of long-term care, hospital admissions, and sometimes contacts of cases. As a less vulnerable population, asymptomatic school children were unlikely to be tested for COVID-19, and K-12 students may represent an understudied population. Our analysis estimates underreporting for the NL provincial testing system, the prevalence and distribution of Omicron variant cases among school students, and the percentage of infections that were asymptomatic for school students that reported positive RAT results.

## 2 Methods

### 2.1 Survey

Parents and guardians of students in grades K-12 that had completed at least one rapid test on January 22 or January 25 were given the opportunity to answer a web survey to report the test results of their household (Health Research Ethics Board, Newfoundland and Labrador, Reference number: 2022.013). Participation was voluntary, and a consent was required before the survey questions were released. Parents and guardians were told that providing the results of the antigen tests would help to understand COVID-19 prevalence and underreporting in NL.

The survey was advertised through broadcast media (2 radio morning shows covering eastern Newfoundland, 2 radio morning shows covering central and western Newfoundland, and Labrador, and 2 evening television news shows covering NL), and on social media (Facebook and Twitter). All principals of private schools, and principals of elementary, junior high, and high schools in the NL English School District were emailed requesting that survey participation details be provided to parents and guardians. All Indigenous groups in the province were emailed information describing how to participate in the study. Exceptions were Innu Nation and Sheshatshiu Innu First Nation School, which returned to school later, and requested that their students complete the RATs on different dates.

The survey consisted of four questions, taking approximately 5 minutes to complete (see Appendix A). Parents and guardians were asked to provide the following information: (1) the first three digits of their postal code, corresponding to the Forward Sortation Area (e.g., A1A), (2) their Regional Health Authority (i.e., Eastern Health, Central Health, Western Health or Labrador-Grenfell Health), the number of rapid tests from their household completed on (3) January 22 and (4) January 25, indicating how many rapid test were negative, positive symptomatic or positive asymptomatic, and specifying whether the students were in grades K-6 or 7-12.

The survey was completed by a total of 1,278 households, where 52% of the households counted more than one student. A total of 2,055 test results were reported (with mostly two test results per student reported), out of an estimated 59,452 students returning to school, which indicates participation of ∼3.5%.

### 2.2 Test accuracy: Sensitivity, specificity, and confidence intervals

The observed number of positive test results *N* ^+^ is the sum of observed positive test results from infected individuals and false positive test results from uninfected individuals, such that:

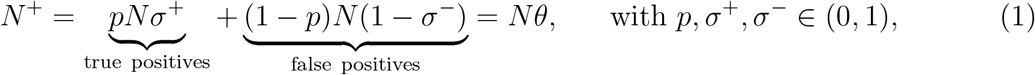

where *p* is the true proportion of infected individuals, *N* is the total number of tests, *θ* is the probability of an individual of testing positive, and *σ*^+^ and *σ*^−^ are sensitivity (i.e., the probability of testing positive if infected) and specificity (i.e., the probability of testing negative if uninfected) respectively.

By rearranging equation (1), we obtain an estimator 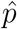 for the true proportion of K-12 students infected with COVID-19:

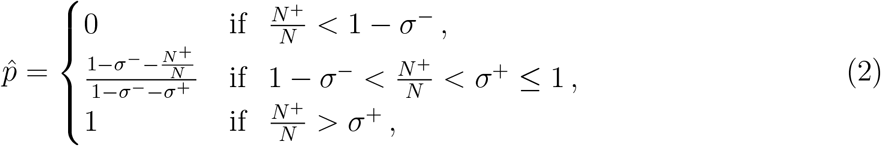

and the estimator 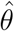 for the probability of testing positive:

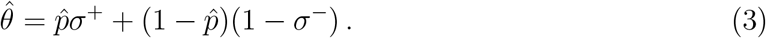

Notice that *N* ^+^ ∼ Bin(*N, θ*). Therefore, Bin 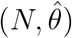 can be resampled to obtain a parametric bootstrap confidence interval estimate.

Sensitivity is estimated as *σ*^+^ = 0.9044. This estimate is based on sensitivity values at different viral loads, and on estimates of viral load during infection [38]. Specificity was assumed to be *σ*^−^ = 0.994, based on the study of Parvu et al. [51]. Testing positive if uninfected is very unlikely (with a mean of 6 out of every 1,000 tests completed), while testing negative if infected can occur with a mean of 1 out of every 10 cases. A complete derivation of the sensitivity and specificity estimates is provided in Appendix C.

The observed number of positive asymptomatic cases includes true positive asymptomatic cases and false positive asymptomatic cases, which could be false positive cases (with very low probability, as discussed above [51]) or positive symptomatic cases falsely reported as asymptomatic. We cannot estimate the proportion of symptomatic cases which may be falsely reported as positive asymptomatic, as this is based on participants’ self-assessment. Therefore, our analysis of asymptomatic cases (Table 2) will be based on the raw reported cases, for which no confidence intervals can be provided.

### 2.3 Data analysis

Anonymized survey results and the code used for the analysis is publicly available at https://github.com/nanomaria/RAT_january. Each row of the data corresponds to the reporting of a single household, where column entries correspond to the number of positive tests (distinguishing between symptomatic and asymptomatic cases), and negative tests in grades K-6 and grades 7-12.

Our analysis provides insights into:

- The rates of underreporting of COVID-19 cases (Omicron variant, BA.1/BA.2 subvariant) in NL at the population level, and at the household level (section 2.3.1).
- The proportion of positive tests occurring in elementary schools (grades K-6) and in junior high and high schools (grades 7-12), and the corresponding proportions of asymptomatic cases (section 2.3.2).
- The spatial distribution of positive households in the province (section 2.3.3).

Test accuracy was taken into account by considering test sensitivity and specificity as described in section 2.2. Data were analysed using the programming language R and the Postal Code^*OM*^ Conversion File (PCCF) [23].

#### 2.3.1 Underreporting

To gain insights into COVID-19 underreporting, we compare estimates of the percentage of positive tests among K-12 students (obtained using the survey-reported rapid antigen test results) with provincial case counts (Appendix D, Fig. D.1). Provincial case counts were based on the Public Service Advisory COVID-19 announcements from the Department and Health and Community Services, which reported the daily number of new cases [11].

In NL, publicly available daily age-structured provincial case counts ended on January 17, 2022, after which only the total number of new cases was provided. By considering age-structured active cases reported till January 17 we derive the percentage of active cases among the younger age group, consisting of individuals aged 0-19 years old (Appendix D, Fig. D.2). We use this percentage to obtain an estimate for reported COVID-19 prevalence among the 0-19 age group when the rapid antigen testing occurred on January 22 and January 25. The procedure is explained in detail in Appendix D. We estimate that 0.49% of the NL population, and 0.45% of youth of age 0-19 years (averaged across January 20 to January 27, 2022) was reported infected with COVID-19. Finally, we use these estimates to quantify the reported household positivity, estimated to be 1.2%. In section 2.4, we discuss how reported COVID-19 prevalence compares with the estimated percent positivity in K-12 students and prevalence of COVID-19 in households, derived from the rapid antigen testing results.

#### 2.3.2 Analysis of positive cases

The total number of positive tests is based on the number of positive tests on January 25 and the number of positive tests on January 22 that have not subsequently been reported on January 25. We define negative tests as the number of negative tests on January 25. This decision was made because parents and guardians were instructed by public health officials to not carry out a second test if the first test was positive, and we decided this after noting that 69 households (out of 1,278) reported different entries on the first and the second testing date (see Appendix B for a complete overview). Positive cases are reported at provincial level, and were divided into symptomatic and asymptomatic cases. The proportion of reported positive cases in elementary (grades K-6) and junior high and high schools (grades 7-12) was also reported.

#### 2.3.3 Spatial distribution of cases

We define positive households as households reporting at least one positive result on either January 22, or January 25. The percentage of positive households was computed at the level of the RHA, and for each FSAs, as described in section 2.2. We perform Moran’s I statistics [48] to investigate the correlation between spatial proximity and COVID-19 prevalence rates in different FSAs.

## Results

### 2.4 Underreporting

When considering the survey-reported RAT results for K-12 students, we estimate that 5.0% (3.8%, 6.5%) of households were positive for COVID-19. When considering the provincial COVID-19 data, we estimate that 1.2% of all households were positive for COVID-19, if we assume that only one test per household was reported in a single day. When comparing our estimates with the provincial estimates we determine that the number of underreported positive households is 4.3 (3.1-5.3) times higher than the counts reported by the NL testing system.

The RAT results that we collected at the individual level indicate a percent positivity of 3.7% (2.9%, 4.7%) among children and youth (Table 1). Provincial reporting was lower, at 0.45% (Appendix D) indicating that on average only one out of every 8.4 (6.4, 10.4) infections has been reported, although we note that this calculation overlooks that infections spread more readily to other household members than to members of the wider community. As discussed previously, we choose to use household positivity (instead of individual positivity) as a more appropriate measure of prevalence given that public health guidelines in place from January 24, 2022 recommended self-testing for household members, with these results not reported in the provincial case counts, and given Omicron’s high transmissibility within households [25].

**Table 1:** RAT results at the provincial level and at the level of the four regional health authorities of NL.

| Region | Total reported positive tests | Total tests | % estimated true positives (95% CI) | Total reported positive households | Total reporting households | % estimated positive households (95% CI) |
| --- | --- | --- | --- | --- | --- | --- |
| Newfoundland and Labrador | 82 | 2055 | 3.7% (2.9%, 4.7%) | 66 | 1278 | 5.0% (3.8%, 6.5%) |
| Eastern Health (EH) | 61 | 1648 | 3.5% (2.5%, 4.5%) | 46 | 1019 | 4.4% (3.0%, 5.8%) |
| Central Health (CH) | 5 | 105 | 4.6% (3.9%, 9.9%) | 5 | 63 | 8.2% (1.1%, 17.0%) |
| Western Health (WH) | 11 | 221 | 4.9% (1.9%, 8.4%) | 10 | 143 | 7.1% (2.4%, 11.8%) |
| Labrador-Grenfell Health (LG) | 5 | 81 | 6.2% (0.7%, 13.1%) | 5 | 53 | 9.8% (1.4%, 18.2%) |

### 2.5 Analysis of positive cases

A total of 82 out of 2055 tests were reported positive, giving an estimate of the true prevalence as 3.7% (2.9%, 4.7%) (Table 2). A larger proportion of these positive tests, namely 62.9% (44.3%, 83.0%), was found in elementary school students, while the remaining 37.1% (22.7%, 52.9%) was estimated in junior high and high school students (grades 7-12). More than half of the cases were reported as asymptomatic (59.8%), with no significant difference in the proportion of asymptomatic cases in grades K-6 and in grades 7-12 (i.e., 60.8% and 58.1% respectively).

**Table 2:** RATs results and estimates of Omicron variant positivity and percent asymptomatic infections in elementary schools (grades K-6) and in junior high and high schools (grades 7-12) in NL.

| Definition | Results |
| --- | --- |
| Total reported positives | 82 |
| Total reported tests | 2055 |
| % estimated true positives | 3.8% (2.9%, 4.7%) |
| Total reported positives (grades K-6) | 51 |
| Total reported positives (grades 7-12) | 31 |
| Total reported (grades K-6) | 1192 |
| Total reported (grades 7-12) | 863 |
| Positives in grades K-6 (% of total estimated true positives) | 62.9% (44.3%, 83.0%) |
| Positives in grades 7-12 (% of total estimated true positives) | 37.1% (22.7%, 52.9%) |
| Total reported asymptomatic | 49 |
| Total reported asymptomatic (grades K-6) | 31 |
| Total reported asymptomatic (grades 7-12) | 18 |
| Asymptomatic (% of total reported positives) | 59.8% |
| Asymptomatic (% of reported positives in grades K-6) | 60.8% |
| Asymptomatic (% of reported positives in grades 7-12) | 58.1% |

### 2.6 Spatial distribution of cases

A total of 66 out of 1,278 households reported at least one positive test on January 22 or January 25, with corresponding household positivity of 5.0% (3.8%, 6.5%). Reports of positive tests were distributed across all four RHAs (Fig. 1). All RHAs reported household positivity larger than 10% in one or more FSAs, as well as low or zero positive tests in other FSAs. FSAs will not be identified, because we do not have consent from the participants to release this information. The population size, area and population density of each RHA is provided in Fig. 1. The total number of households reporting is provided in Table 1.

**Figure 1:**
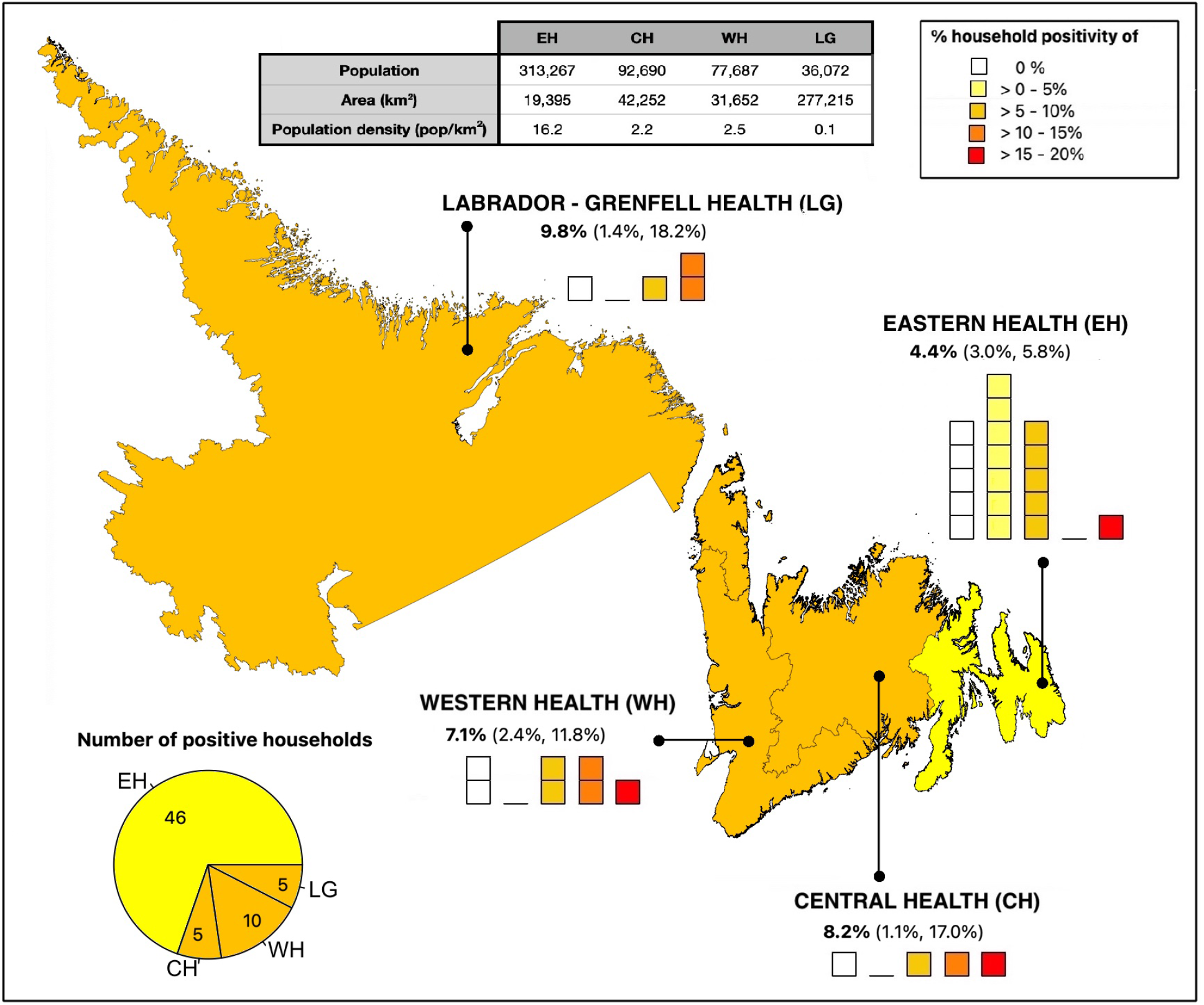
Map of Newfoundland and Labrador (NL), divided by Regional Health Authorities (RHAs), from left to right, Labrador-Grenfell Health (LG), Western Health (WH), Central Health (CH), and Eastern Health (EH). The household positivity (i.e., the proportion of households which reported positive test results from K-12 students) reported by each of its Forward Sortation Areas (FSAs) is shown for each RHA, where each square corresponds to a single FSA within the RHA and the color of the square represents the reported household positivity. We include only results of FSAs for which test results of students of six or more households have been reported. Percent values represent the percentage of positive households in each region, with 95% confidence intervals. The pie chart represents the number of reported positive households for each health region as a fraction of the total number of positive households in the province. The population, area, and population density of each RHA are provided in the table in the top of the figure.

Participants from Eastern Health (EH), as the smallest but most populated health region of the province, reported results for 17 out of 18 FSAs. This region reported 46/1019 positive households (out of 66 total positive households in the whole province), but lower COVID-19 prevalence rates with respect to other health regions, with the percentage of positive households being 4.4% (3.0%, 5.8%). Participants from Central Health (CH) reported 5/63 positive households and a household positivity of 8.2% (1.1%, 17.0%), based on the RAT results of 4 out of 7 FSAs. Participants from Western Health (WH) reported results from 7 out of 7 FSAs, with 10/143 positive households and household positivity of 7.1% (2.4%, 11.8%). Participants from Labrador and Grenfell Health (LG) reported 5/53 positive households, with a household positivity estimated as 9.8% (1.4%, 18.2%). The number of FSAs reporting low versus high percentage household positivity for each RHA is provided in Fig. 1. Because of lower reporting rates and possible sampling biases, there is high uncertainty associated with household positivity in the regions of LG, CH, and WH, and, more generally, with prevalence at the FSAs level.

To understand whether geographically close FSAs reported similar results, we perform Moran’s I statistics to measure the correlation between percent household positivity and proximity. We obtain a Moran’s I coefficient of -0.08, and a p-value of 0.35, indicating no correlation between spatial proximity and COVID-19 prevalence rates among FSAs.

## Discussion

COVID-19 underreporting has been a major challenge for pandemic monitoring and response planning. These difficulties have increased with the establishment of the highly transmissible Omicron variant [30, 53], and due to the expanded use of rapid tests, which are not officially reported in some jurisdictions [30, 64]. Underreporting rates may have been particularly high among children and youth, given their relatively low risk of experiencing severe outcomes [24]. In NL, public health officials recommended that all K-12 students complete RATs on January 22 and 25, 2022. We conducted an online survey where parents and guardians of K-12 students could report these RAT results. Self-administered rapid tests were not reported in the NL provincial case counts, and characteristics of the NL population eligible for testing under the provincial system [11], were very different than the characteristics of the population that completed the RATs on January 22 and 25, 2022. We estimated that only 1 out of every 8.4 (6.4-10.4) cases occurring in children and youth, or 1 out of every 4.3 (3.1-5.3) positive households, was reported by provincial case counts.

The COVID-19 Immunity Taskforce uses serological analysis of blood donations to estimate the percentage of provincial populations that have been infected with SARS-CoV-2 [1]. When interpreted relative to the number of cases reported by the NL testing system, these serology data imply that from January 15 to February 12, 2022, 1 in every 2.3 cases were reported (Table E.1). For comparison, in other Canadian provinces from January 15 to February 12, 2022, the underreporting ratio ranged from 1 in every 17.2 cases reported (British Columbia) to all cases reported (Prince Edward Island, Table E.1). Underreporting ratios are generally highest in children [56]. The underreporting ratio that we estimate from our study of rapid antigen testing in K-12 students is broadly consistent with COVID-19 Immunity Taskforce data. Eligibility for testing, such that the results of the testing could be reported in the provincial case counts, was relatively unrestricted in NL at the time of our study, while in all other provinces except for Prince Edward Island, most individuals were ineligible for testing under the provincial systems due to age restrictions on eligibility (see Appendix E for complete details).

We find that most of the positive cases occurred in elementary schools (62.9%, 95% CI: 44.3%, 83.0%), while previous research found higher COVID-19 prevalence in junior high and high schools (secondary schools) relative to elementary schools (primary schools; [34, 41, 42]), presumably, due to student cohorting. Elementary school students tend to remain with the same classmates throughout the day, while older students have different classmates in different classes. However, for the RAT results collected in our study, testing was conducted after schools had been closed for five weeks. Therefore, student cohorting or other public health measures aimed to reduce COVID-19 spread in schools did not impact our results. Potentially, a major factor influencing results is the vaccination status of the students. Vaccination rates for 5-11 year olds in NL where the highest in Canada, with 75% having received one date of vaccine on January 19, 2022 [17]. However, youth aged 12 years or older became eligible for vaccination starting May 23, 2021, while children aged 5 and above became eligible for vaccination only on November 23, 2021. At the time of our study, on January 22, 2022, nearly all junior and high school-aged youths were fully vaccinated (96.7% of NL residents aged 12-17 years), while nearly all elementary school-aged children had not completed a full vaccination series (only 3.3% of NL residents aged 5-11 years had completed a full vaccination series [13]).

Whether children and youth are more susceptible than adults to SARS-CoV-2 infection has been a matter of debate [60]. Understanding the role that children play in the transmission of the virus is key to inform public health policies for the implementation of non-pharmaceutical interventions, such as school closures. Given the consequences of school closures on mental and social health [59, 63], it is important to understand the effect that closing schools has on COVID-19 transmission. Understanding the role of school children in SARS-CoV-2 spread may also help inform vaccine prioritization strategies. Possible vaccination strategies include prioritizing essential workers (e.g., teachers, or other workers with a large number of social contacts), which would reduce transmission and the total number of infections [49].

We estimate that 59.8% of the positive tests were asymptomatic, where asymptomatic rates were similar among elementary school students (60.8%) and students in junior high and high schools (58.1%). Previous studies have reported asymptomatic rates associated with the Omicron variant to be between 32% and 44% [55], where asymptomatic rates tend to be higher in younger age groups [29, 47, 55]. Our high asymptomatic rates could be due to reporting errors. For example, in some instances (see Appendix B) participants reported asymptomatic infection on February 22, and symptomatic infection on February 25, which indicates a possible confusion between asymptomatic and pre-symptomatic infections. Infections asymptomatic at the time of the testing, but with symptoms appearing some days later, should have been reported as symptomatic, but may have been reported as asymptomatic instead, which would lead to an overestimation of the percentage of asymptomatic infections. On the other hand, the survey was conducted two weeks after the RATs were taken, such that participants were given enough time to realize whether symptoms occurred during the infectious period, and correctly report whether infections were symptomatic or not. It could be possible that asymptomatic rates in children and youth are effectively high or that the estimate is unreliable due to low sample size.

The RAT survey results also allow us to investigate the spatial distribution of COVID-19 cases. We find high heterogeneity in the percentage of positive cases reported across the province, and no relationship between regional proximity and COVID-19 prevalence. Although a positive correlation between COVID-19 prevalence and population density may have been expected [57, 62], we find that Eastern Health, the RHA with the highest population density, reported the lowest infection prevalence. Due to our small sample size, we can not determine whether the low counts registered for Eastern Health are an artifact of higher reporting rates, and whether using a finer spatial scale or having a larger dataset for certain FSAs could have revealed more insights into the spatial pattern of cases. Previous studies have also found marked heterogeneity in the spatial distribution of COVID-19 cases [31, 32], where household size, rather than population density, has been recognized to be a better indicator of COVID-19 hotspots [45, 46].

Given the low participation rate in the survey (3.5%) and small sample sizes, and given that participation in the survey was voluntarily, our results may suffer from sample selection biases, and should be interpreted with caution. It may be that those households with positive tests were more likely to report results, which may have inflated positive case counts in comparison to provincial estimates. Additionally, different social and psychological stresses may have resulted in certain social groups (such as pro- or anti-vaccine groups) being more likely to report results than others, leading to additional biases. Finally, sources of bias occur in the provincial testing system also, for example higher testing rates of vulnerable individuals, hospital admissions, and long-term care residents, many of whom are elderly. Nonetheless, altogether our study provides a complete analysis of the survey data to understand possible patterns of Omicron prevalence among children and youth, a currently understudied population.

## Data Availability

All data produced in the present work are contained in the manuscript

## Acknowledgement

We wish to thank the Nunatsiavut Government, NunatuKavut Community Council, and the Mik’maq (Northern Peninsula Band, Burgeo Band, St. Georges Band, Benoit First Nation, and Qalipu First Nation) all of whom were gracious and responsive to our requests for consultation regarding this research. We thank Katie Winters for translating the research summary into Labrador Inuttitut. We thank David Champredon, Caroline Colijn, Monika Dutt, David J.D. Earn, Marie-Josée Fortin, Jane Heffernan, Sarah P. Otto and Robert Way for comments on the analysis methodology. We gratefully acknowledge logistical support from Yolanda Wiersma, George Adu-Boahen, Francis Anokye, and Ashley Locke. We acknowledge the Newfoundland and Labrador broadcast media for their role in having this study gain public awareness, and in particular, Ramraajh Sharvendiran.

## Appendix

### Appendix A: Survey

#### COMPLETE 1 SURVEY FOR ALL K-12 STUDENTS IN YOUR HOUSEHOLD

##### CONSENT TO PARTICIPATE IN THE RESEARCH STUDY

###### ELIGIBILITY

a. You are a parent or guardian of K-12 student(s) in Newfoundland and Labrador, and
b. K-12 student(s) in your household completed <u>at least one rapid antigen test</u> on January 22 and/or January 25.

**Do you consent to participate in the research study?** (YES/NO)

1. **Enter the <u>first 3-digits</u> of your postal code, i.e. A1A**:

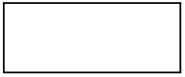
2. **What is your Regional Health Authority?**
  ∘Eastern Health
  ∘Central Health
  ∘Western Health
  ∘Labrador Grenfell
  At the recommendation of the Department of Health and Community Services, on January 22 and 25 many students in Newfoundland and Labrador completed COVID-19 rapid antigen tests prior to returning to school. For question 3. (below) the determination of ‘no symptoms’ or ‘with symptoms’ refers to the period January 19 to 29. For a definition of symptoms, please see: https://www.gov.nl.ca/covid-19/public-health-guidance/covid-19/symptoms
3. **For the FIRST rapid test(s) completed on January 22, the number of rapid tests from your household:**

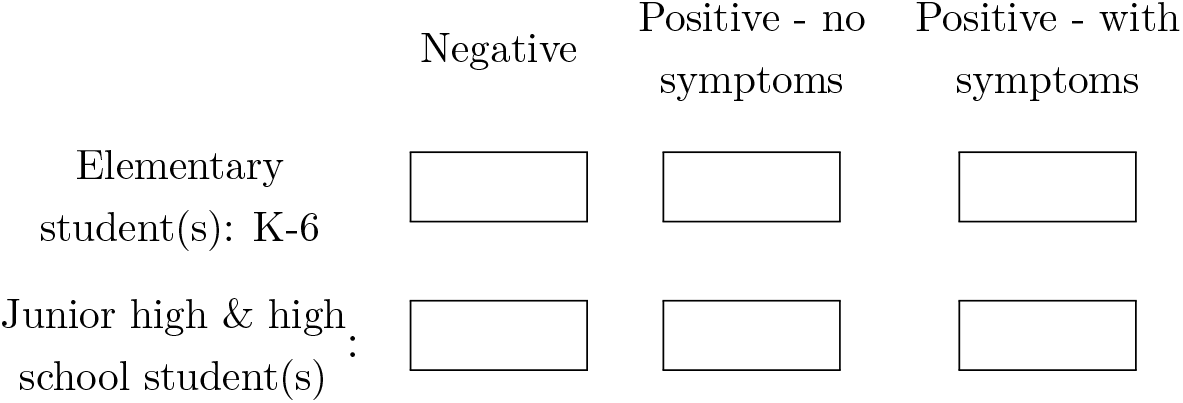 For question 4. (below) the determination of ‘no symptoms’ or ‘with symptoms’ refers to the period January 22 to February 1. For a definition of symptoms please see: https://www.gov.nl.ca/covid-19/public-Health-guidance/covid-19/symptoms/
4. **For the SECOND rapid test(s) completed on January 25, the number of rapid tests from your household:**

**4. For the SECOND rapid test(s) completed on January 25, the number of rapid tests from your household:**
|  | Negative | Positive - no symptoms | Positive - with symptoms |
| --- | --- | --- | --- |
| Elementary student(s): K-6 |  |  |  |
| Junior high & high school student(s) |  |  |  |

### Appendix B

To understand how testing results differed between January 22 and January 25 we examine the reporting of 69 households out of 1,278 that reported different results on the two dates. We found the following noteworthy changes between the two dates:

- *Positive to no report*: Parents and guardians were instructed to not conduct a second test on January 25, if the first test on January 22 were positive. We counted 9 households that reported positive cases on January 22 and no testing on January 25, presumably because of following the provincial guidelines.
- *Positive asymptomatic to symptomatic*: Four households reported a change from positive asymptomatic cases on January 22, to positive symptomatic cases on January 25, possibly indicating that cases reported as asymptomatic were instead pre-symptomatic cases. The distinction between symptomatic and asymptomatic cases was based on participants’ self-assessment, and it is possible that pre-symptomatic cases may have been reported as asymptomatic instead.
- *Negative to positive*: We found that 29 households reported negative cases on January 22, and positive cases on January 25 (16 asymptomatic and 13 symptomatic), likely indicating latent infections, where the viral load was not high enough to be detected by the first test, but successively increased enough to lead to a positive result three days later.
- *Positive to negative*: We found that 3 households reported cases changing from positive to negative between January 22 and 25 (one household reported one case changing from asymptomatic positive to negative, one household reported one case changing from symptomatic positive to negative, and one household reported one case changing from asymptomatic positive to negative and one case changing from symptomatic positive to negative). This change could be due to recovery (particularly for the household reporting a symptomatic positive case and for the household reporting two positive tests, both changing to negative three days later), or to a false positive result, although multiple studies have shown that the probability of testing positive if uninfected is very low (e.g., 0.006% [51]). We consider positive households as households that reported at least one positive result on January 22 or 25, and therefore according to our definition these households were considered positive.
- *Different total number of reported tests* : We found that 36 households reported a different total number of tests performed on January 22 and January 25. This difference may be due to students being in different households on the two dates, especially given that the first day was to be completed on a Saturday, while the second test was to be completed on a Tuesday, the morning before schools reopening.

A detailed overview of this data is provided in the publicly accessible database at https://github.com/nanomaria/RAT_january.

### Appendix C: Sensitivity and specificity

**Table C.1:** Data used to derive the averaged sensitivity of repeated testing. Estimations of viral load are given in log units (e.g., 5 log units = 100,000 copies/mL). Sensitivity at different viral loads during infection (rounded to the nearest integer log unit) are obtained from an Ontario Science Table publication [38]. Sensitivity for repeated testing is the probability of obtaining a positive test if infected on either January 22 or January 25 (see Eq. (4)). Total average sensitivity is the averaged sensitivity of obtaining a positive test on January 22 or on January 25, assuming uniform distribution of exposure times. We consider sensitivities between day 0 and day 14 after exposure.

| Days from exposure (test 1) | Days from exposure (test 2) | Viral load (test 1) [log<sub>10</sub> Units] | Viral load (test 2) [log<sub>10</sub> Units] | Sensitivity (test 1) | Sensitivity (test 2) | Sensitivity for repeated testing |
| --- | --- | --- | --- | --- | --- | --- |
| — | 0 | 0 | 3.0 | — | 16.9% | 16.9% |
| — | 1 | 0 | 5.0 | — | 64.0% | 64.0% |
| — | 2 | 0 | 7.0 | — | 98.0% | 98.0% |
| 0 | 3 | 3.0 | 9.0 | 16.9% | 99.3%* | 99.42% |
| 1 | 4 | 5.0 | 9.0 | 64.0% | 99.3%* | 99.75% |
| 2 | 5 | 7.0 | 8.2 | 98.0% | 99.3% | 99.986% |
| 3 | 6 | 9.0 | 7.6 | 99.3%* | 99.3% | 99.995% |
| 4 | 7 | 9.0 | 7.0 | 99.3%* | 98.0% | 99.986% |
| 5 | 8 | 8.2 | 6.6 | 99.3% | 98.0% | 99.986% |
| 6 | 9 | 7.6 | 6.2 | 99.3% | 94.1% | 99.959% |
| 7 | 10 | 6.0 | 6.0 | 98.0% | 94.1% | 99.88% |
| 8 | 11 | 6.6 | 5.6 | 98.0% | 94.1% | 99.88% |
| 9 | 12 | 6.2 | 5.4 | 94.1% | 84.3% | 99.07% |
| 10 | 13 | 6.0 | 5.2 | 94.1% | 84.3% | 99.07% |
| 11 | 14 | 5.6 | 5.0 | 94.1% | 84.3% | 99.07% |
| 12 | — | 5.4 | — | 84.3% | — | 84.3% |
| 13 | — | 5.2 | — | 84.3% | — | 84.3% |
| 14 | — | 5.0 | — | 84.3% | — | 84.3% |
| * We choose sensitivity for a viral load of 9 log<sub>10</sub> Units (not provided in Juni et al, 2021) to be equal to the sensitivity of a viral load of 8 log<sub>10</sub> Units |  |  |  |  | **Averaged sensitivity** | 90.44% |

#### Sensitivity

To determine the overall probability of testing positive if infected on either January 22 or January 25, we consider sensitivity values at different viral loads to a rapid antigen test reported by the Ontario Science Table ([38], based on data from [52]).

Additionally, we consider estimates of viral load during infection to determine test sensitivity from day 0 to day 14 after exposure (from Jüni et al. [38], based on data from Li et al. [43] and Kang et al. [39]). As viral load curves for Omicron (with various vaccination statuses) are currently not known, we use viral load curves derived for the Delta variant, therefore assuming similarities in the analytical sensitivity of RATs to the Delta and Omicron variants [28]. Note that sensitivities can vary by manufacturers [54].

To estimate sensitivity of repeated testing, we derive the probability of testing positive if infected when a second test is performed three days after the first one (e.g., the first test is performed on day 2 after exposure, while the second test is performed on day 5 after exposure), taking into account differences in viral load at different days after exposure. Estimates are provided in Table C.1. For each of the dates considered, we have

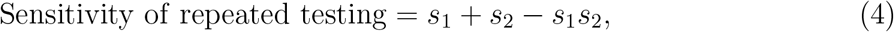

where *s*_1_ is the sensitivity of test 1 (i.e., the test performed on January 22) and *s*_2_ is the sensitivity of test 2 (i.e., the test performed on January 25). The total averaged sensitivity *σ*^+^ = 0.9044 is obtained by averaging the sensitivity of repeated testing when considering uniform distribution of exposure times.

#### Specificity

Specificity does not depend on viral load and is assumed to be the same on January 22 and on January 25. As we consider positive cases as individuals testing positive on either January 22 or January 25, the specificity of repeated testing equals the probability of single testing. We choose *σ*^−^ = 0.994%, based on the meta-analysis of Parvu et al. [51], indicating that it is highly unlikely to test positive if uninfected.

### Appendix D: Provincial case counts and underreporting

#### Reported positivity

Fig. D.1 shows the number of active cases based on the Public Service Advisory COVID-19 updates that report the number of new cases [11], as a percentage of the population of Newfoundland and Labrador (estimated to be 510,550 [2]), and assuming a recovery time of 7 days. For data reported by the government of NL, the definition of active cases was changed from 10 to 7 days for the data reported from January 18, 2022 onward. As we are interested in comparing testing results with the number of active cases in the province around these dates (recall that rapid test were performed on January 22 and January 25), we prefer to derive the number of active cases from new cases such that we can use a consistent recovery time throughout the time period of interest.

The mean percentage of people in NL reported as active COVID-19 cases from January 20 to January 27 was estimated to be 0.49%. After January 17, case reporting by age group was no longer provided by the province. To estimate COVID-19 prevalence in K-12 students we multiply this estimate by the percentage of cases occurring in individuals of age 0-19, averaged between January 1 and January 14 (Fig. D.2), and weighted by the total population aged 0-19 years old (estimated to be 93,910 [2]). We estimate reported COVID-19 prevalence in youth at the time of the rapid antigen testing to be 0.45%.

#### Reported household positivity

We consider the reported COVID-19 prevalence in NL (i.e., 0.49%, as derived above) to estimate household positivity. Given that we can not differentiate between positive tests reported from households with or without students in grades K-12, we estimate COVID-19 prevalence at provincial level, where the total number of households in Newfoundland and Labrador is 223,255 [2].

Before January 24, 2022, rapid antigen testing was recommended for household close contacts of someone with COVID-19 who has symptoms [11]. Close contacts testing positive on a rapid antigen test, were recommended to book a Polymerase Chain Reaction (PCR) test. From January 24, testing was recommended only for non-household contacts, with specific vaccination status [11]. If we assume that only one test per household was reported in a single day, we obtain a household positivity of 0.49% *×* population (NL) *÷* 223, 255 = 1.2%.

**Figure D.1:**
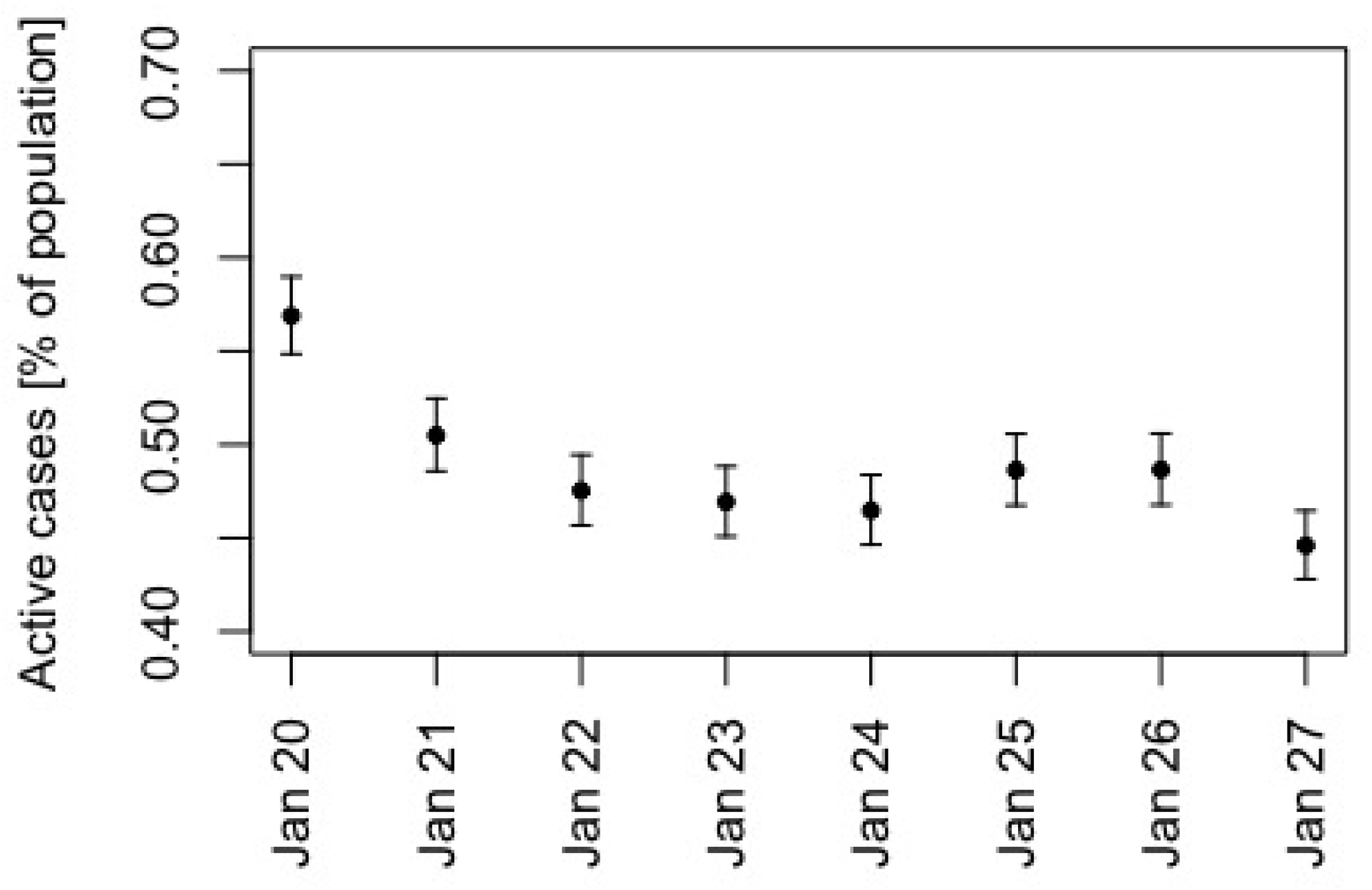
Active cases as a percentage of the population of Newfoundland and Labrador, between January 20 and January 27, 2022, with corresponding 95% confidence intervals (Jeffrey intervals). Estimates are derived from the Public Service Advisory updates reporting new cases of COVID-19 [11] and assuming a recovery time of 7 days.

**Figure D.2:**
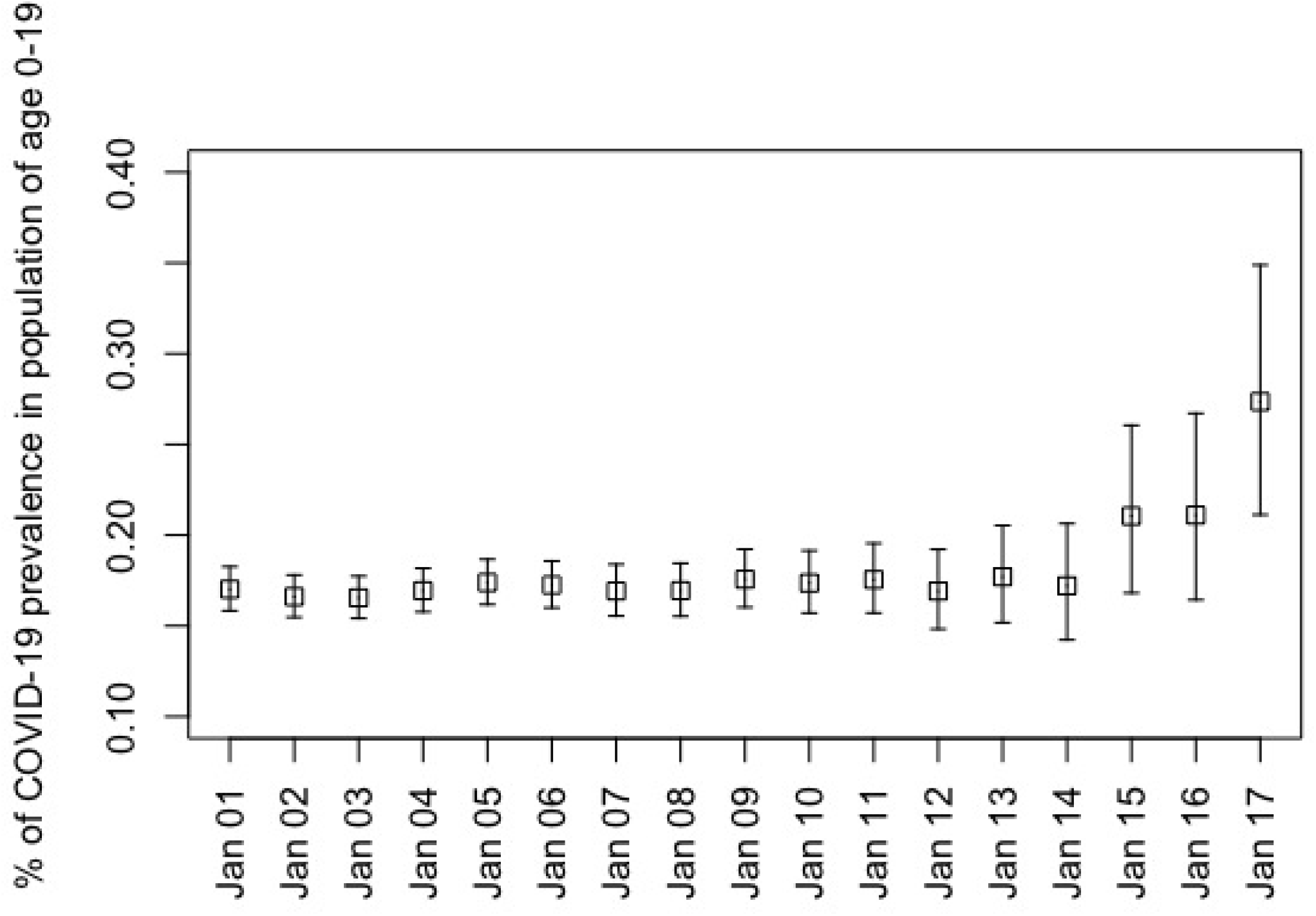
Percentage of reported COVID-19 cases occurring in individuals of age 0-19, between January 1 and January 17, 2022, with corresponding 95% confidence intervals (Jeffrey intervals). Estimates are based on the Public Service Advisory updates for COVID-19 in NL [11], and on census data [2]. For our computations, we consider only the average of the daily reported percentages from January 1 to January 14, as the percentage of reported COVID-19 cases occurring in youth seems to be fairly constant during this period of time. After January 14, we can observe a trend of a higher percentage of cases in youth, although uncertainty for these dates is higher. On January 17, 2022 the province stopped reporting age-structured case counts, and this trend can not be confirmed by available data.

### Appendix E: Estimating underreporting from COVID-19 Immunity Task-force serology data

Provincial population sizes are from 2021 census estimates [2]. For January 15 and February 12, 2022 (a 28-day period), cumulative reported cases were downloaded from the Public Health Agency of Canada website [12]. The cumulative percentage of the population infected are estimates reported by the COVID-19 Immunity Taskforce [1] derived from serology of Canadian Blood Services and Héma-Québec donations. Seroconversion occurs 9-12 days after symptom onset [44], and serology data for comparison to the reported cases is considered from January 28 to February 25, 2022 (a 28-day period). The number of underreported cases per reported case was calculated as the difference in the percentage of the population infected as estimated by the serology, divided by the difference in the percentage of the population reported as infected.

**Table E.1:** Underreporting of COVID-19 cases for January-February, 2022, calculated from the COVID-19 Immunity Taskforce serology data

|  | Population size | Cumulative reported cases to Jan 15, 2022 | Cumulative reported cases per capita to Jan 15, 2022 | Cumulative reported cases to Feb 12, 2022 | Cumulative reported cases per capita to Feb 12, 2022 | Seropositivity to Jan 28, 2022 (95% CI) | Seropositivity to Feb 25, 2022 (95% CI) | Estimated unreported cases per reported case |
| --- | --- | --- | --- | --- | --- | --- | --- | --- |
| British Columbia | 5,000,879 | 295,904 | 5.9% | 339,736 | 6.8% | 15.9%<br>(13.4, 18.5) | 31.4%<br>(28.3, 34.5) | 17.2 |
| Alberta | 4,262,635 | 439,320 | 10.3% | 514,251 | 12.1% | 20.5%<br>(17.7, 23.5) | 36.1%<br>(32.6, 39.6) | 8.7 |
| Saskatchewan | 1,132,505 | 98,699 | 8.7% | 125,756 | 11.1% | 14.7%<br>(12.0, 17.4) | 28.0%<br>(24.3, 31.6) | 5.5 |
| Manitoba | 1,342,153 | 107,838 | 8.0% | 125,844 | 9.4% | 22.6%<br>(19.5, 25.8) | 31.9%<br>(28.1, 35.6) | 6.6 |
| Ontario | 14,223,942 | 937,636 | 6.6% | 1,070,455 | 7.5% | 16.4%<br>(13.8, 19.1) | 26.2%<br>(23.0, 29.5) | 10.9 |
| Quebec | 8,501,833 | 806,920 | 9.5% | 899,260 | 10.6% | 14.3%<br>(8.9, 20.2) | 28.8%<br>(22.3, 35.8) | 13.2 |
| New Brunswick | 775,610 | 22,359 | 2.9% | 32,610 | 4.2% | 3.7%<br>(2.4, 5.4) | 11.0%<br>(8.2, 14.3) | 5.6 |
| Nova Scotia | 969,383 | 30,101 | 3.1% | 42,230 | 4.4% | 5.4%<br>(3.5, 7.4) | 15.8%<br>(12.3, 19.6) | 8.0 |
| Prince Edward Island | 154,331 | 4,118 | 2.7% | 10,105 | 6.6% | 2.2%<br>(1.1, 3.9) | 6.0%<br>(3.2, 9.8) | 1.0 |
| Newfoundland and Labrador | 510,550 | 12,406 | 2.4% | 19,954 | 3.9% | 2.7%<br>(1.3, 4.6) | 6.1%<br>(3.6, 9.2) | 2.3 |

On December 24 2021, most residents of Nova Scotia became ineligible to be tested under the provincial testing system, and were instead directed to complete rapid tests, tests that would identify infections, but where positive results were not reported in the provincial case counts [9]. All other provinces, except for NL and Prince Edward Island, had similar testing eligibility requirements, i.e., testing eligibility under the provincial systems was restricted to high risk groups (i.e., hospital patients), and individuals in contact with high risk groups (i.e., patient-facing health care workers) [10], and with the results of testing completed outside the provincial system not included in provincial case counts.

Prince Edward Island introduced an appointment-based system for COVID-19 testing on June 8, 2022, but unlike all other Canadian provinces, never introduced restricted eligibility requirements for provincial testing.

Only on March 17, 2022, did NL introduce restrictions to testing eligibility [6] that were similar to the restricted eligibility enacted in all other provinces, except Prince Edward Island, by early January 2022. On January 4, 2022, testing eligibility in NL excluded symptomatic contacts of cases [5]. On January 24, 2022, testing eligibility excluded household members of cases and symptomatic close contacts of cases, however, asymptomatic close contacts of cases (non-household members) and symptomatic individuals that were not a close contact were still recommended for testing under the provincial system [4], such that testing eligibility in NL was relatively unrestricted until March 17, 2022, when compared to all other provinces except Prince Edward Island.

## References

[1] Results: Blood Donation Organizations. https://www.covid19immunitytaskforce.ca/results-blood-donation-organizations/. Accessed 2022-08-01.

[2] Statistics canada. 2022. (table). Census Profile. 2021 Census of Population. Statistics Canada Catalogue no. 98-316-X2021001. Ottawa. Released April 27, 2022. https://www12.statcan.gc.ca/census-recensement/2021/dp-pd/prof/index.cfm?Lang=E (accessed May 27, 2022).

[3] Public Advisory: Update on COVID-19 in Newfoundland and Labrador. January 7, 2022. https://www.gov.nl.ca/releases/2022/health/0107n04/.

[4] Public Advisory: Update on COVID-19 in Newfoundland and Labrador; Province to Remain in Modified Alert Level 4. https://www.gov.nl.ca/releases/2022/health/0124n05/,.

[5] Public Advisory: Update on COVID-19 in Newfoundland and Labrador. January 3, 2022. https://www.assembly.nl.ca/legislation/sr/statutes/a04-2.htm,.

[6] Public Advisory: Revised Eligibility Criteria for PCR Testing and Direction for Cases of COVID-19. March 17, 2022. https://www.gov.nl.ca/releases/2022/health/0317n11/.

[7] Report to the House of Assembly on the COVID-19 Public Health Emergency. https://www.assembly.nl.ca/business/electronicdocuments/ReporttoHOACOVID-19PublicHealthEmergency2022.pdf.

[8] Report to the house of assembly on the covid-19 public health emergency. https://www.assembly.nl.ca/business/electronicdocuments/ReporttoHOACOVID-19PublicHealthEmergency2022.pdf (accessed Dec 6, 2022).

[9] Changes in COVID-19 Testing and Case Management. December 24, 2021. https://novascotia.ca/news/release/?id=20211224001. Accessed 2022-08-04.

[10] Updated Eligibility for PCR Testing and Case and Contact Management Guidance in Ontario. https://news.ontario.ca/en/backgrounder/1001387/updated-eligibility-for-pcr-testing-and-case-and-contact-management-guidance-in-ontario.

[11] Public advisory: Update on covid-19 in newfoundland and labrador. https://www.gov.nl.ca/releases/covid-19-news/. Accessed 2022-05-26.

[12] COVID-19 Epidemiology update. https://health-infobase.canada.ca/covid-19/. Accessed 2022-08-01,.

[13] COVID-19 vaccination in Canada. https://health-infobase.canada.ca/covid-19/vaccination-coverage/,.

[14] Rapid testing program for students and staff at schools. newfoundland and labrador. https://www.gov.nl.ca/covid-19/schools-children/school-rapid-testing-program/. Accessed 2022-05-04.

[15] Classification of Omicron (B.1.1.529): SARS-CoV-2 Variant of Concern. https://www.who.int/news/item/26-11-2021-classification-of-omicron-(b.1.1.529)-sars-cov-2-variant-of-concern. Accessed November 26, 2021.

[16] Cbc news. canada’s first cases of the omicron coronavirus variant confirmed in ottawa. https://www.cbc.ca/news/politics/omicron-variant-canada-travellers-1.6265927. Accessed June 7, 2022.

[17] How did Newfoundland manage to vaccinate 75 per cent of 5-11-year-olds? https://www.macleans.ca/news/how-did-newfoundland-manage-to-vaccinate-75-percent-of-5-11-year-olds/.

[18] Updates on COVID-19 Variants of Concern (VOC). https://nccid.ca/covid-19-variants/. Accessed March 7, 2022.

[19] N.L. students, staff return to the classroom after COVID-19 delay. https://www.cbc.ca/news/canada/newfoundland-labrador/nl-return-to-school-jan-2022-1.6325652. Accessed Jun 16, 2022.

[20] Public Advisory: K-12 Schools Closing Two Days Early for Christmas Break. https://www.gov.nl.ca/releases/2021/education/1219n03/, accessed June 7, 2022.

[21] COVID-19 vaccination in Canada. https://health-infobase.canada.ca/covid-19/vaccination-coverage/, accessed May 31, 2022.

[22] An act respecting the attainment of the age of majority. https://www.gov.nl.ca/releases/2022/health/0103n02/, 1995.

[23] Postal Code^OM^ Conversion File (PCCF). 2017.

[24] Atheer Atiroğlu, Ahmed Atiroğlu, Münteha Özsoy, Vesen Atiroğlu, and Mahmut Özacar. COVID-19 in adults and children, symptoms and treatment. Biointerface Research in Applied Chemistry, 12(2):1735–1748, 2022.

[25] Julia M Baker, Jasmine Y Nakayama, Michelle O’Hegarty, Andrea McGowan, Richard A Teran, Stephen M Bart, Katie Mosack, Nicole Roberts, Brooke Campos, Alina Paegle, et al. SARS-CoV-2 B. 1.1. 529 (Omicron) variant transmission within households—four US jurisdictions, November 2021–February 2022. Morbidity and Mortality Weekly Report, 71(9):341, 2022.

[26] Michael G Baker, Nick Wilson, and Tony Blakely. Elimination could be the optimal response strategy for COVID-19 and other emerging pandemic diseases. BMJ, 371, 2020.

[27] Matthew J Binnicker. Challenges and controversies to testing for COVID-19. Journal of clinical microbiology, 58(11):e01695–20, 2020.

[28] Joshua Deerain, Julian Druce, Thomas Tran, Mitchell Batty, Yano Yoga, Michael Fennell, Dominic E Dwyer, Jen Kok, and Deborah A Williamson. Assessment of the analytical sensitivity of ten lateral flow devices against the SARS-CoV-2 omicron variant. Journal of Clinical Microbiology, pages jcm–02479, 2021.

[29] Carolina Diamandis, Jonathan Feldman, and Adrian Tudor. Asymptomatic Covid-19: a major source of infection at the onset of an Omicron storm. Authorea Preprints, 2021.

[30] Ahmed Elbanna. Estimation of the Ascertainment Bias in Covid Case Detection During the Omicron Wave. medRxiv, 2022.

[31] Yongjiu Feng, Qingmei Li, Xiaohua Tong, Rong Wang, Shuting Zhai, Chen Gao, Zhenkun Lei, Shurui Chen, Yilun Zhou, Jiafeng Wang, et al. Spatiotemporal spread pattern of the COVID-19 cases in China. PLoS One, 15(12):e0244351, 2020.

[32] Claudio Fronterre, Jonathan M Read, Barry Rowlingson, Simon Alderton, Jessica Bridgen, Peter J Diggle, and Chris P Jewell. COVID-19 in England: spatial patterns and regional outbreaks. medRxiv, 2020.

[33] Zhiru Gao, Yinghui Xu, Chao Sun, Xu Wang, Ye Guo, Shi Qiu, and Kewei Ma. A systematic review of asymptomatic infections with COVID-19. Journal of Microbiology, Immunology and Infection, 54(1):12–16, 2021.

[34] Deepti Gurdasani, Nisreen A Alwan, Trisha Greenhalgh, Zoë Hyde, Luke Johnson, Martin McKee, Susan Michie, Kimberly A Prather, Sarah D Rasmussen, Stephen Reicher, et al. School reopening without robust COVID-19 mitigation risks accelerating the pandemic. The Lancet, 397(10280):1177–1178, 2021.

[35] Anita E Heywood and C Raina Macintyre. Elimination of COVID-19: what would it look like and is it possible? The Lancet Infectious Diseases, 20(9):1005–1007, 2020.

[36] Amy Hurford, Maria M Martignoni, Jesus Concepcion Loredo-Osti, Francis Anokye, Julien Arino, Bilal Saleh Husain, Brian Gaas, and James Watmough. Pandemic modelling for regions implementing an elimination strategy. medRxiv, 2022.

[37] Nahla Khamis Ibrahim. Epidemiologic surveillance for controlling covid-19 pandemic: types, challenges and implications. Journal of infection and public health, 13(11):1630– 1638, 2020.

[38] P Jüni, S Baert, P Bobos, et al. Rapid antigen tests for voluntary screen testing. Science Briefs of the Ontario COVID-19 Science Advisory Table, 2:52, 2021.

[39] Min Kang, Hualei Xin, Jun Yuan, Sheikh Taslim Ali, Zimian Liang, Jiayi Zhang, Ting Hu, Eric Lau, Yingtao Zhang, Meng Zhang, et al. Transmission dynamics and epidemiological characteristics of Delta variant infections in China. Medrxiv, 2021.

[40] Kai Kupferschmidt. New mutations raise specter of ‘immune escape’. Science, 371 (6527):329–330, 2021. doi: 10.1126/science.371.6527.329. URL https://www.science.org/doi/abs/10.1126/science.371.6527.329.

[41] Elisabetta Larosa, Olivera Djuric, Mariateresa Cassinadri, Silvia Cilloni, Eufemia Bisaccia, Massimo Vicentini, Francesco Venturelli, Paolo Giorgi Rossi, Patrizio Pezzotti, Emanuela Bedeschi, et al. Secondary transmission of COVID-19 in preschool and school settings in northern Italy after their reopening in September 2020: a population-based study. Eurosurveillance, 25(49):2001911, 2020.

[42] Eva Leidman, Lindsey M Duca, John D Omura, Krista Proia, James W Stephens, and Erin K Sauber-Schatz. COVID-19 trends among persons aged 0–24 years—United States, March 1–December 12, 2020. Morbidity and Mortality Weekly Report, 70(3):88, 2021.

[43] Baisheng Li, Aiping Deng, Kuibiao Li, Yao Hu, Zhencui Li, Yaling Shi, Qianling Xiong, Zhe Liu, Qianfang Guo, Lirong Zou, et al. Viral infection and transmission in a large, well-traced outbreak caused by the SARS-CoV-2 Delta variant. Nature Communications, 13(1):1–9, 2022.

[44] Zheng SF Su YY Li ZY Liu W Yu F Ge SX Zou QD Yuan Q Lin S Hong CM Yao XY Zhang XJ Wu DH Zhou GL Hou WH Li TT Zhang YL Zhang SY Fan J Zhang J Xia NS Chen Y. Lou B, Li TD. Serology characteristics of SARS-CoV-2 infection since exposure and post symptom onset. Vaccines, 56(2):2000763, 2020.

[45] Andrew R Maroko, Denis Nash, and Brian T Pavilonis. COVID-19 and inequity: a comparative spatial analysis of New York City and Chicago hot spots. Journal of Urban Health, 97(4):461–470, 2020.

[46] Christopher A Martin, David R Jenkins, Jatinder S Minhas, Laura J Gray, Julian Tang, Caroline Williams, Shirley Sze, Daniel Pan, William Jones, Raman Verma, et al. Sociodemographic heterogeneity in the prevalence of COVID-19 during lockdown is associated with ethnicity and household size: Results from an observational cohort study. EClinicalMedicine, 25:100466, 2020.

[47] Seyed Mohammad Miri, Farshid Noorbakhsh, Seyed Reza Mohebbi, and Amir Ghaemi. Higher prevalence of asymptomatic or mild COVID-19 in children, claims and clues. Journal of Medical Virology, 2020.

[48] Patrick AP Moran. Notes on continuous stochastic phenomena. Biometrika, 37(1/2): 17–23, 1950.

[49] Nicola Mulberry, Paul Tupper, Erin Kirwin, Christopher McCabe, and Caroline Colijn. Vaccine rollout strategies: The case for vaccinating essential workers early. PLOS Global Public Health, 1(10):e0000020, 2021.

[50] Miguel I Paredes, Stephanie M Lunn, Michael Famulare, Lauren A Frisbie, Ian Painter, Roy Burstein, Pavitra Roychoudhury, Hong Xie, Shah A Mohamed Bakhash, Ricardo Perez, et al. Associations between SARS-CoV-2 variants and risk of COVID-19 hospitalization among confirmed cases in Washington State: a retrospective cohort study. Medrxiv, 2021.

[51] Valentin Parvu, Devin S Gary, Joseph Mann, Yu-Chih Lin, Dorsey Mills, Lauren Cooper, Jeffrey C Andrews, Yukari C Manabe, Andrew Pekosz, and Charles K Cooper. Factors that Influence the Reported Sensitivity of Rapid Antigen Testing for SARS-CoV-2. Frontiers in Microbiology, page 2611, 2021.

[52] Tim Peto, Dominic Affron, Babak Afrough, Anita Agasu, Mark Ainsworth, Alison Allanson, Katherine Allen, Collette Allen, Lorraine Archer, Natasha Ashbridge, et al. COVID-19: Rapid antigen detection for SARS-CoV-2 by lateral flow assay: A national systematic evaluation of sensitivity and specificity for mass-testing. EClinicalMedicine, 36:100924, 2021.

[53] Carolina Ribeiro Xavier, Rafael Sachetto Oliveira, Vinícius da Fonseca Vieira, Marcelo Lobosco, and Rodrigo Weber dos Santos. Characterisation of Omicron Variant during COVID-19 Pandemic and the Impact of Vaccination, Transmission Rate, Mortality, and Reinfection in South Africa, Germany, and Brazil. BioTech, 11(2):12, 2022.

[54] John G Routsias, Maria Mavrouli, Panagiota Tsoplou, Kyriaki Dioikitopoulou, and Athanasios Tsakris. Diagnostic performance of rapid antigen tests (RATs) for SARS-CoV-2 and their efficacy in monitoring the infectiousness of COVID-19 patients. Scientific reports, 11(1):1–9, 2021.

[55] Weijing Shang, Liangyu Kang, Guiying Cao, Yaping Wang, Peng Gao, Jue Liu, and Min Liu. Percentage of asymptomatic infections among SARS-CoV-2 Omicron variant-positive individuals: a systematic review and meta-analysis. Vaccines, 10(7):1049, 2022.

[56] Danuta M Skowronski, Samantha E Kaweski, Michael A Irvine, Shinhye Kim, Erica SY Chuang, Suzana Sabaiduc, Mieke Fraser, Romina C Reyes, Bonnie Henry, Paul N Levett, et al. Serial cross-sectional estimation of vaccine and infection-induced SARS-CoV-2 sero-prevalence in children and adults, British Columbia, Canada: March 2020 to August 2022. medRxiv, 2022.

[57] Karla Therese L Sy, Laura F White, and Brooke E Nichols. Population density and basic reproductive number of COVID-19 across United States counties. PloS One, 16 (4):e0249271, 2021.

[58] Vikram Thakur and Radha Kanta Ratho. OMICRON (B. 1.1. 529): A new SARS-CoV-2 Variant of Concern mounting worldwide fear. Journal of Medical Virology, 2021.

[59] Russell Viner, Simon Russell, Rosella Saulle, Helen Croker, Claire Stansfeld, Jessica Packer, Dasha Nicholls, Anne-Lise Goddings, Chris Bonell, Lee Hudson, et al. Impacts of school closures on physical and mental health of children and young people: a systematic review. MedRxiv, 2021.

[60] Russell M Viner, Oliver T Mytton, Chris Bonell, GJ Melendez-Torres, Joseph Ward, Lee Hudson, Claire Waddington, James Thomas, Simon Russell, Fiona Van Der Klis, et al. Susceptibility to SARS-CoV-2 infection among children and adolescents compared with adults: a systematic review and meta-analysis. JAMA pediatrics, 175(2):143–156, 2021.

[61] J Weinberg. Surveillance and control of infectious diseases at local, national and international levels. Clinical Microbiology and Infection, 11:12–14, 2005.

[62] David WS Wong and Yun Li. Spreading of COVID-19: Density matters. Plos One, 15 (12):e0242398, 2020.

[63] Joseph T Wu, Shujiang Mei, Sihui Luo, Kathy Leung, D. Liu, Qiuying Lv, Jian Liu, Yuan Li, Kiesha Prem, Mark Jit, et al. A global assessment of the impact of school closure in reducing COVID-19 spread. Philosophical Transactions of the Royal Society A, 380(2214):20210124, 2022.

[64] Pei Yuan, Elena Aruffo, Yi Tan, Liu Yang, Nicholas H Ogden, Aamir Fazil, and Huaiping Zhu. Projections of the transmission of the Omicron variant for Toronto, Ontario, and Canada using surveillance data following recent changes in testing policies. Infectious Disease Modelling, 7(2):83–93, 2022.

